# RS-fMRI Evidence of Left Frontal Lobe Developmental Deviation as a Potential Pathognomonic Feature of Autism Spectrum Disorder

**DOI:** 10.1101/2025.04.01.25325010

**Authors:** Tien-Wen Lee

## Abstract

Autism spectrum disorder (ASD) is one of the most prevalent developmental disorders. This study utilized 3-Tesla resting-state fMRI data analyzed with the functional parcellation algorithm MOSI (Modularity Analysis and Similarity Measurements) to investigate cortical functional organization in ASD. Sixty individuals with ASD and sixty healthy controls were recruited, with no significant differences in age and gender distribution. The MOSI-derived metrics were compared using independent two-sample t-tests. The findings revealed a significant reduction in the functional volume of the left frontal lobe, a region critical for language and social processing. This reduction appears to be accompanied by compensatory expansion in other brain regions, suggesting a reallocation of neural resources that may contribute to ASD heterogeneity. These results support the notion of left frontal lobe developmental deviation (LFDD) as a parsimonious neural mechanism underlying core ASD features. The accountability of LFDD in various cognitive, symptomatic, and behavioral characteristics of ASD is briefly discussed, along with its implications for male predominance and evolutionary relevance. Overall, these findings provide a novel brain science perspective that moves beyond traditional psychological frameworks in explaining major psychiatric disorders.

**Lay summary:** This study reveals that individuals with autism spectrum disorder (ASD) exhibit a significant reduction in the functional volume of the left frontal lobe, a brain region crucial for language and social skills. This alteration may lead to compensatory changes in other brain areas, contributing to the diversity of ASD traits.

## Introduction

Autistic spectrum disorder (ASD) is a developmental disorder that is characterized by difficulties in social interaction and social communication, as well as repetitive, restricted, and inflexible patterns of behavior, interests, and activities, along with atypical sensory processing (American Psychiatric Association, 2013). From a brain science perspective, several theories of ASD have been proposed, each addressing a particular cognitive domain or characteristic manifestation, such as theory of mind (ToM), executive function, central coherence, and “extreme male brain” (Baron-Cohen, 2002; Baron-Cohen, 1991; Silva et al., 2013). Recent research has emphasized neural connectivity abnormalities in ASD (Goodwill et al., 2023). The complexity inherent in ASD (and other major psychiatric conditions) has been a significant challenge in constructing comprehensive mechanistic models.

In contrast to the neural node- or circuitry-based approach, which largely draws from psychological or computational perspectives, the author proposed major depressive disorder (MDD) as a compartment-level disorder (Lee & Xue, 2018), which not only possessed higher accountability and was recently empirically verified (Lee, 2025a). “Compartment” here refers to the frontoparietal mantle or limbic system as neural entity (Lee et al., 2014), not confined to a specific region, network, or psychological construct. The theory formulation of MDD exemplifies the process of “dialectic neuroscience,” which seeks to integrate diverse evidence into a parsimonious disease mechanism (Lee, 2025b). The treatment of neural compartments requires functional parcellation of the cortex as a pre-processing step, achieved through a relatively novel algorithm—modular analysis and similarity measurements (MOSI) (Lee & Tramontano, 2021). Then, akin to the case of MDD illustrated above, is it possible to move beyond psychological approaches and employ a similar framework to decipher ASD?

Previous theories of ASD, although each supported by empirical evidence, have limited explanatory power. Take ToM as an example: while it provides an appealing construct for examining social dysfunction in ASD, it fails to account for abnormalities in executive control, restricted interests, and central coherence. Furthermore, ToM deficits are not unique to ASD but are also observed in other psychiatric conditions, such as schizophrenia (Bora et al., 2009). Taking a more holistic view, language deficits, communication difficulties, executive dysfunction, intellectual disability, repetitive behaviors, restricted interests, and features associated with the extreme male brain hypothesis may be more cohesively understood through the concept of left frontal lobe dysfunction as a unifying factor—or more precisely, developmental abnormalities of the left frontal lobe (elaborated in the **Discussion**).

This study applied MOSI to resting-state functional magnetic resonance imaging (rsfMRI) data to investigate the brain lobes (frontal, temporal, limbic, and parietal) as the subjects of analysis. The resulting lobar metrics of ASD patients will be statistically compared with those of age- and gender-matched neurotypical controls.

## Methods

### Subjects, MRI data, and preprocessing

MRI data from 120 participants, all aged over 18 to minimize variance due to the maturation process, were obtained from the Autism Brain Imaging Data Exchange (ABIDE) (Di Martino et al., 2014). The cohort included 60 individuals diagnosed with ASD and 60 neurotypical controls (HC), with written consent acquired from all participants. Functional and structural MRI (sMRI) images of the entire brain were acquired using 3.0 Tesla MRI scanners, with parameters set to TR = 1.5s, voxel size = 3×3×4 mm^3^, and a total of 200 volumes.

The echo planar imaging (EPI) data was processed using the Analysis of Functional NeuroImages (AFNI) software package (Cox, 1996). The preprocessing pipeline for rsfMRI included steps such as despiking, slice-time correction, motion realignment, T1 anatomical registration, spatial smoothing, and bandpass filtering in the range of 0.01–0.10 Hz. To ensure magnetization equilibrium, the first six scans (9 seconds) were discarded. A spatial smoothing kernel of 6 mm was applied to the EPI data. The FreeSurfer software was used for segmenting gray and white matter from T1-weighted structural MRI (sMRI) data, as well as for parcellating the cortical mantle into 68 regions of interest (ROIs) based on the Desikan-Killiany Atlas (Dale et al., 1999; Desikan et al., 2006; Fischl et al., 1999), which served as the initial partition for the subsequent MOSI analysis. Following the analytic pipeline described by Jo et al. (Jo et al., 2010; Lee et al., 2014; Xue et al., 2014), 12 movement parameters, along with signals from white matter and ventricles, were included in the regression model (Jo et al., 2010). Additionally, second-order polynomials were applied to account for baseline drift. Functional connectivity (FC) was calculated by computing the pairwise Pearson correlation coefficient (CC) between the temporal traces of any paired voxels or the mean temporal traces of partitioned clusters by MOSI.

### MOSI analysis

The connectivity map constructed from Pearson correlation coefficients (CCs) serves as the basis for modular analysis. In this platform, modules—also referred to as communities—are sets of nodes within a network that exhibit higher intra-group connectivity compared to inter-group connectivity, as determined by CCs. MOSI integrates both modular analysis and similarity measurements, iteratively dividing modules into smaller sub-modules and merging similar sub-modules into larger ones. The parcellation process in MOSI adheres strictly to two principles: spatial proximity and functional similarity. Adjacent voxels with comparable neural activity are grouped together, aligning with the fundamental logic of functional parcellation (FP) in cortical signal processing. This iterative split-merge process continues until the system reaches convergence. The current implementation of MOSI employs the Louvain community detection algorithm, where the granularity of modular partitioning is adjustable via the Gamma parameter. Higher Gamma values yield a greater number of modules, refining the resolution of the analysis (Lee & Tramontano, 2021). The author examined 12 different resolutions, with Gamma values ranging from 0.30 to 0.85. Each segmented module was assigned to a specific brain lobe based on majority voting when it spanned two or more lobes. The MOSI-derived metrics, including module count and size, were compared between the ASD and HC groups using independent two-sample t-tests. Since the outputs from different Gamma values are not entirely independent, applying a Bonferroni correction would be overly conservative and could increase the risk of false negatives. Therefore, statistical thresholds were set at p-values < 0.01 and < 0.005 to represent different levels of stringency.

## Results

The mean age of the HC and ASD groups was 21.0 years (SD = 4.2) and 20.9 years (SD = 4.0), respectively, with no significant difference between the groups (t(118) = −0.1323, P = 0.895). Both groups had identical gender distributions, each comprising 57 male participants.

In alignment with the multi-resolution feature of MOSI, higher Gamma values led to an increase in the number of clusters while reducing the voxel count per cluster. In the HC group, the cortical module count reached 98 at Gamma 0.30 and 233 at Gamma 0.85. Similarly, in the ASD group, the module count was 100 at Gamma 0.30 and 239 at Gamma 0.85. The initial partition, based on the Desikan-Killiany Atlas, comprised 68 modules.

For the between-group comparisons, independent t-tests indicated no significant differences in the partition numbers derived from MOSI across the 12 resolutions (Gamma values) between the ASD and HC groups. Likewise, the voxel counts of the four cortical lobes did not show any group differences in anatomical volume. Notably, however, ASD participants exhibited a statistically significant reduction in the “functional volume” of the left frontal lobe (LFL), particularly at lower Gamma values. Given that MOSI-derived modules may span across two anatomical lobes, majority voting was employed to assign each module to a specific brain lobe. Accordingly, functional volumes are not equal to anatomical volumes that are determined by landmarks but were calculated based on the total voxel count of modules classified within each brain lobe. A summary of the anatomical and functional volume findings is presented in Table 1.

**Table 1.** Statistical Comparisons of Functional Volumes in the Left Frontal Lobe.

| Gamma | ASD |  | HC |  | t-scores | p-values |
| --- | --- | --- | --- | --- | --- | --- |
|  | mean | std | mean | std |  |  |
| 0.30 | 2923.3 | 655.4 | 3291.1 | 627.2 | -3.14 | 0.002** |
| 0.35 | 2903.2 | 664.4 | 3298.8 | 647.9 | -3.30 | 0.001** |
| 0.40 | 2910.8 | 712.4 | 3249.7 | 626.9 | -2.77 | 0.007* |
| 0.45 | 2878.8 | 694.1 | 3284.1 | 642.3 | -3.32 | 0.001** |
| 0.50 | 2807.7 | 735.6 | 3283.1 | 662.2 | -3.72 | 0.0003** |
| 0.55 | 2809.7 | 768.6 | 3210.3 | 681.5 | -3.02 | 0.003** |
| 0.60 | 2675.5 | 773.0 | 3146.9 | 666.0 | -3.58 | 0.001** |
| 0.65 | 2628.4 | 736.1 | 2995.0 | 686.8 | -2.82 | 0.006* |
| 0.70 | 2597.5 | 762.2 | 2897.2 | 789.5 | -2.11 | 0.037 |
| 0.75 | 2477.6 | 664.4 | 2782.4 | 767.1 | -2.33 | 0.022 |
| 0.80 | 2415.8 | 682.7 | 2635.0 | 728.1 | -1.70 | 0.092 |
| 0.85 | 2438.6 | 617.8 | 2530.4 | 677.4 | -0.78 | 0.439 |

The contents are voxel number. \* and \*\* indicate p-values less than 0.01 and 0.005, respectively.

Further investigation revealed that the significance cannot be solely attributed to one factor, such as the parietal lobe enlargement at the expense of the functional volume reduction in the left frontal lobe (statistics not significant). Rather, the reduction appears to be a synergistic outcome resulting from the inherent trend of smaller anatomical volume in the LFL, combined with the influence of other brain regions. For completeness, exploratory analyses, such as regional power, integrative graph indices, and power-connectivity relationships were carried out, but all showed negative results (data thus not present).

## Discussion

Elucidating the neural mechanisms underlying ASD remains a persistent challenge in brain science. Through the novel functional parcellation scheme MOSI, left frontal lobe abnormality emerged as the only significant finding. A key advantage of MOSI lies in its individualized and multi-resolution approach—the former addressing ASD heterogeneity, while the latter enables examining ASD pathology at an optimal scale (resolution) (Lee & Tramontano, 2021). Building on MOSI, the concept of functional volume is introduced to distinguish it from anatomical volume. The reduction in functional volume of the left frontal lobe indicates that other brain regions may expand, highlighting the underdevelopment of the left frontal lobe. This suggests a potential shift in neural resource allocation, where compensatory mechanisms in other areas could occur as a result of the developmental deviation in the left frontal lobe, which might be further relevant to some ASD individuals exhibiting exceptional abilities in restricted areas. Notably, at higher resolutions, the partitioned modules become smaller, reducing the likelihood and impact of modules spanning multiple brain lobes—underscoring the value of multi-resolution exploration in psychopathology. Dialectic neuroscience has pursued an integrative approach—synthesizing analyses—to address complex issues (Lee, 2016, 2025b), with accountability playing a central role. In the following discussion, an attempt is made to propose left frontal lobe developmental deviation (LFDD) as a parsimonious neural mechanism to explain the core features of ASD.

First of all, LFDD theory implies that the developmental abnormalities may spare the posterior cortical regions. This argument generally holds from two aspects. Firstly, perceptual issues in ASD primarily involve abnormal sensitivities, rather than a deficit in the ability to discriminate sensory stimuli, which may be influenced by top-down control mechanisms. Some ASD cases may exhibit high-functioning skills in areas such as mathematics (arguably related to parietal region), memory (temporal region), or pattern recognition. Secondly, the cortical development follows a posterior-to-anterior progression, with the perceptual and motor functions maturing first, followed by the frontal lobe, where the left side matures later than the right (Gerván et al., 2017). This developmental sequence aligns with evolutionary patterns, where the expansion of the frontal lobe is considered a later evolutionary event undergoing fewer selection processes, making it more prone to disruptions (Hoffmann, 2013; Pletikos et al., 2014).

LFDD may also account for a broad range of characteristic impairments in ASD, particularly social dysfunction due to cognitive deficits. The left frontal lobe is often associated with higher-order cognitive functions such as language, logic, and analytical thinking. In the context of social communication, it plays a role in tasks involving verbal communication, organizing thoughts, and processing social information in a structured or logical manner. Evidence suggests bilateral hemispheric interaction through mutual inhibition, indicating that LFDD may also impact right frontal functioning, including emotional and social domains. The accountability of LFDD in response inhibition, attention regulation, restricted interest, central coherence, and behavioral symptoms are well supported by brain science literature, but the review is beyond the scope of this short report (Dichter, 2012; Silva et al., 2013). Given the broad connections modulated by frontal lobe, widespread connectivity abnormalities are reasonably expected.

There is emerging speculation that autism reflects an exaggeration of male-typical cognitive patterns. While existing literature, such as that by Baron-Cohen, largely attributes the male predominance in ASD to androgen effects (Baron-Cohen, 2002), this research suggests that deficits in LFL development may also play a critical role, particularly when considering the gender differences in the lateralization of frontal functions. This insight becomes particularly compelling when considering that sex hormones may exert a substantial influence on immature neural tissues during development. As a result, delayed or reduced development in the LFL—critical for language and social processing—could result in prolonged and excessive influence from sex hormones, leading to a more pronounced bias in males.

The lobar-level abnormalities in ASD align with the compartment-level analogs observed in MDD, moving beyond psychological frameworks and interpretations to seek an alternative brain science perspective as the core explanation for major psychiatric disorders. Lobe-level abnormalities occur at a broader, coarser scale, inevitably triggering compensatory and plasticity changes at finer scales to counteract dysfunction, processes that are highly individualized (Kanai & Rees, 2011; Kolb & Gibb, 2014; Poldrack, 2000). Varying degrees of LFDD and various compensatory changes may account for the heterogeneity in ASD, and in some cases, exceptional skills may result from compensatory expansion of non-frontal brain tissues, as implied by this study. As this research focuses solely on the cortex, parallel developmental deviations in subcortical structures cannot be excluded (Persichetti et al., 2025).

## Data Availability

All data produced in the present study are available upon reasonable request to the authors

## Authors Contributions

This report is authored by a single individual.

## Acknowledgments

The author would like to thank Autism Brain Imaging Data Exchange (ABIDE) for their generosity in sharing MRI data. This work was supported by NeuroCognitive Institute (NCI) and NCI Clinical Research Foundation Inc.

## Statements and Declarations

No conflicts of interest to declare.

## Compliance with ethical standards

This research analyzed the databank from a publicly released dataset. The author carried out no animal or human studies for this article.

